# Integrated Bioinformatic Approach to Identify Potential Biomarkers against Idiopathic Pulmonary Fibrosis

**DOI:** 10.1101/2019.12.12.19014746

**Authors:** Md. Asad Ullah, Bishajit Sarkar, Yusha Araf, Md. Nazmul Islam Prottoy, Ananna Saha, Tanjila Jahan, Aisha Siddiqua Boby, Md. Shariful Islam

## Abstract

Idiopathic Pulmonary Fibrosis (IPF) is a chronic and progressive lung disease that leads to gradual decline in lung function. The molecular mechanism and risk factors of this disease are still obscure. Poorly understood etiology of this disease is the major obstacle in the identification of potential biomarkers and drug targets. In this study, microarray gene expression data of normal and IPF patient has been utilized for the statistical analysis of differentially expressed genes (DEGs) with a view to identifying potential molecular signatures using network-based system. Then their functional enrichment analysis revealed their predominant involvement in transcription, protein acetylation, extracellular matrix organization, apoptic process, inflammatory response etc. Protein-Protein Interaction (PPI) network revealed (UBC, PTEN, SOS1, PTK2, FGFR1, YAP1, FOXO1, RACK1, BMP4 and CD44) as hub proteins in IPF. Subsequent regulatory network analysis suggested (E2F1, STAT3, PPARG, MEF2A, FOXC1, GATA3, YY1, GATA2, NFKB1, and FOXL1) as the best regulatory transcriptional signatures and (hsa-mir-155-5p, hsa-mir-16-5p, hsa-mir-17-5p, hsa-mir-19a-3p, hsa-mir-192-5p, hsa-mir-92a-3p, hsa-mir-26b-5p, hsa-mir-335-5p, hsa-mir-124-3p, and hsa-let-7b-5p) as the best post-transcriptional signatures. This study represents proteome and RNA signatures of IPF which might be useful to uphold the present efforts in the discovery of potential biomarkers and treatments of this disease.

## 1. Introduction

Idiopathic Pulmonary Fibrosis (IPF) is a progressive and degenerative interstitial pneumonia that results in the abnormal scarring of the lungs of affected individual (Selman et al., 2001). Historically IPF was thought to be a chronic inflammatory process but there is now growing evidence that IPF response is mediated by abnormally activated alveolar epithelial cells (AECs) which secrete mediators that drive the growth of fibroblast and myofibroblast foci. These foci in turn secrete excessive extracellular matrix which then form abnormal scarring resulting in the alteration of lung architecture (King et al., 2001; Fernandez and Eickelberg, 2012). Previously IPF was accused to lead to the steady and predictable decline in lung function but now there are enough evidences that IPF also leads to the heart failure, pulmonary embolism and acute respiratory deterioration (Collard et al., 2007). The clinical epidemiology of IPF remains poorly understood since the first identification of this disease. However, environmental exposures, genetic factors and tobacco smoking are considered as the potential risk factors of IPF. There are now two approved drugs i.e., Pirfenidone and Nintedanib but unfortunately surgical therapy and lung transplantation are merely accessible to every people (Ley and Collard, 2013; Hughes et al., 2016). Older age with a median of 66 years is the major clinical feature of IPF and in addition, older age has also been shown to exhibit poorer prognosis (Ley et al., 2011). Report suggests that, IPF prevalence in USA varies between 43 and 67 cases per 100,000 population. Moreover, the occurrence and prevalence of IPF increases with age and the incidence in males is higher than females (Nalysnyk et al., 2012).

Microarray technology now allows the easy identification of variance in gene expression which in turn contributes in the identification of potential biomarkers of specific interest (Giltnane and Rimm, 2004; Kerr et al., 2000). Analysis of differential gene expression signatures is crucial in the identification of biomarkers and understanding of lung pathologies in IPF (DePianto et al., 2015; Li et al., 2014). However, although these studies provide fruitful findings but the prediction of actual mechanism of biological condition using the data of differentially expressed genes (DEGs) is often difficult and may come with erroneous interpretation sometime (Crow et al., 2019).

In this study, we have employed integrated network-based strategies to predict potential biomarkers which may contribute in the identification of new drug target, and early diagnosis of IPF. Thereafter we have also tried to discuss the specific roles of identified signatures in IPF (**Figure 1**).

**Figure 1:**
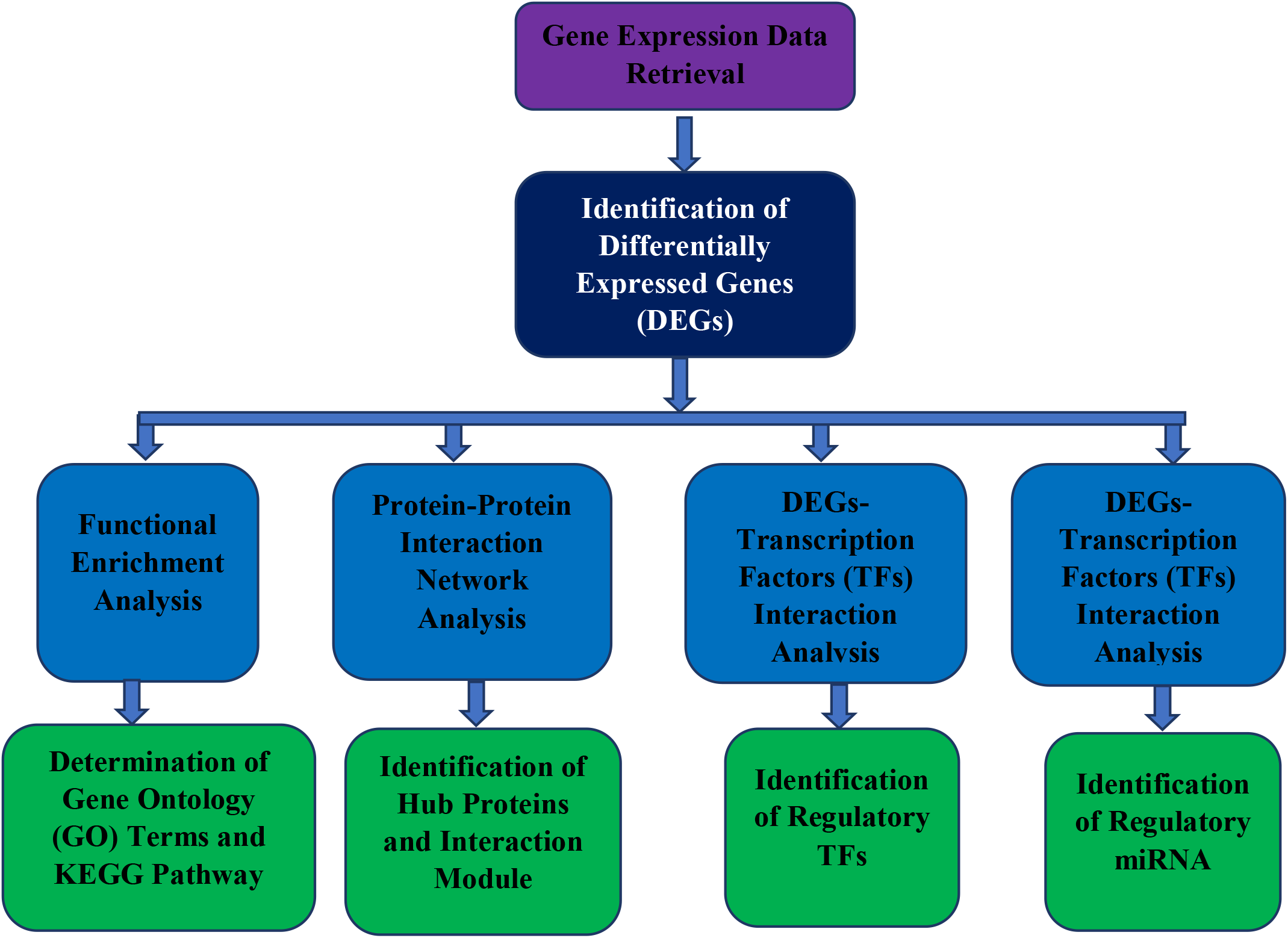
Flowchart of strategies employed in the overall study.

## 2. Materials and Methods

### 2.1. Data Retrieval and Identification of Differentially Expressed Genes

We retrieved GSE24206 microarray data from NCBI-GEO (National Center for Biotechnology Information-Gene Expression Omnibus) database (Meltzer et al., 2011). The dataset comprises the expression profile of lung tissues from different parts of lungs of 5 normal donors and 11 IPF patients. After retrieval, the data was analyzed using GEAP (Gene Expression Analysis Platform) to differentiate the upregulated and downregulated genes (Nunes et al., 2018). Log2 transformation was applied and differentially expressed genes (DEGs) were sorted with adjusted P value<0.01 filter since the lower value corresponds to more accurate prediction.

### 2.2. Functional Enrichment Analysis of DEGs

Both upregulated and downregulated gene sets were analyzed by DAVID (Database for Annotation, Visualization, and Integrated Discovery) (version 6.8) for gene over-representation to elucidate gene ontology (GO) terms and pathways involved with DEGs (Shermanand and Lempicki, 2009). P values were adjusted using the Hochberg and Benjamini test and gene count >2 were set as the cut-off point during the analysis.

### 2.3. Construction of Protein-Protein Interaction Network and Identification of Hub Proteins

STRING database was utilized for the reconstruction of protein-protein interaction (PPI) network with NetworkAnalyst (Szklarczyk et al., 2017; Xia et al., 2015). Topological and expression analysis of the DEGs were performed using NetworkAnalyst. Hub proteins in the generic PPI network with top 10 most connected nodes were identified with cytoHubba plugin using bottleneck interaction matrix on Cytoscape (version 3.7.2) (Chin et al., 2014; Shannon et al., 2003). Top 2 modules in the network was also analyzed using MCODE plugin (Saito et al., 2012).

### 2.4. Identification of Regulatory Molecules

DEGs were searched against JASPAR with the help of NetworkAnalyst to construct transcription factor (TF)-DEGs interaction network (Sandelin et al., 2004). Micro RNA (miRNA)-DEGs interaction network was constructed searching the DEGs against TarBase (Sethupathy et al., 2006). Top 10 interacting TFs and miRNAs were selected and analyzed.

## 3. Result

### 3.1. Transcriptome Signatures

Publicly available GEO data from NCBI was utilized in this study in order to identify differentially expressed genes. A total of 660 DEGs were identified with 276 downregulated genes and 384 upregulated genes within the selected parameters after statistical analysis (**Figure 2**).

**Figure 2:**
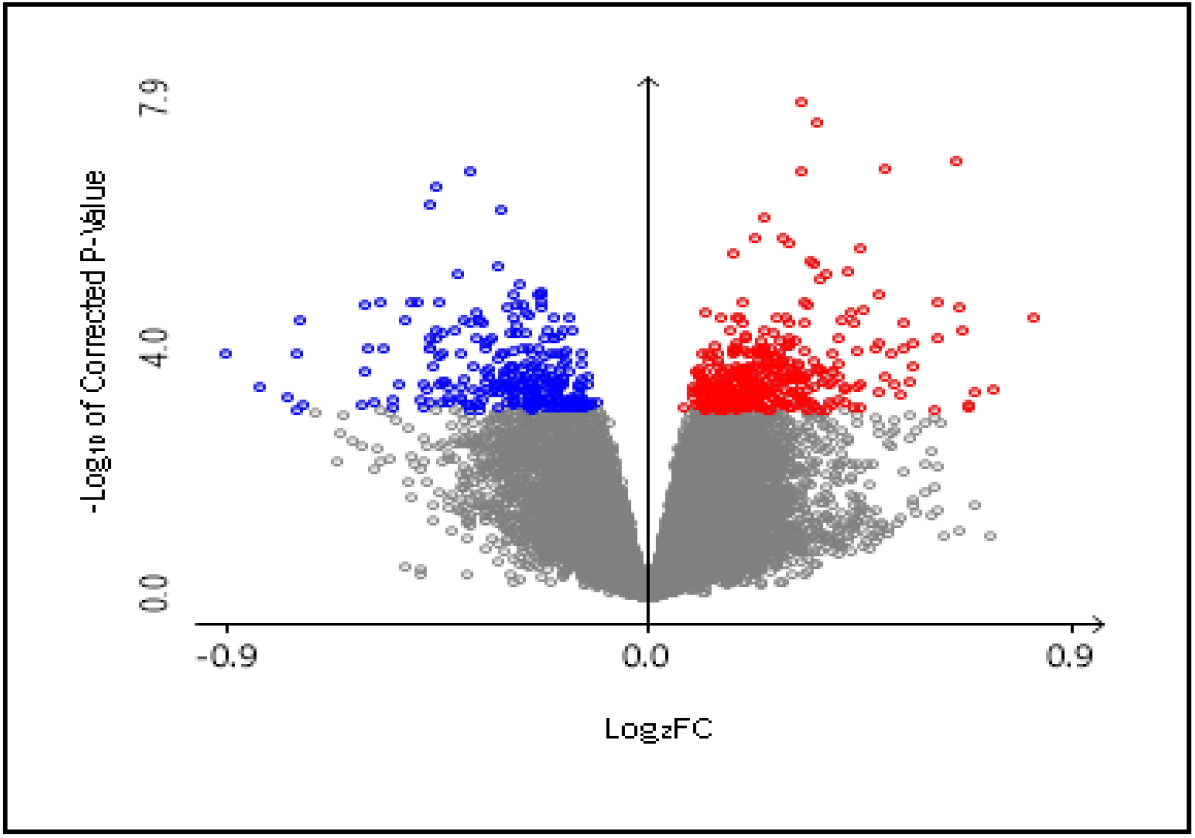
Volcano plot of selected differentially expressed genes (DEGs). Colored (Blue: Down regulated genes; Red: Upregulated genes) DEGs have been selected with adjusted P value > 0.01. filter.

Thereafter the selected genes were analyzed to understand the functional enrichment reflecting significant GO terms and enriched pathway. Top 5 GO terms were retained for both upregulated and downregulated genes (**Table 1**). The DEGs were then subjected to analyze their involvement in biological pathway. Selected genes showed sign of their involvement in KEGG (Kyoto Encyclopedia of Genes and Genomes) pathway (**Figure 3**).

**Table 1:**
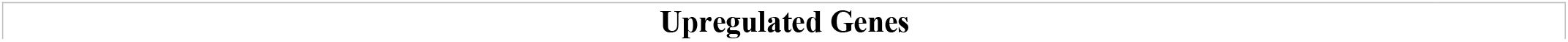
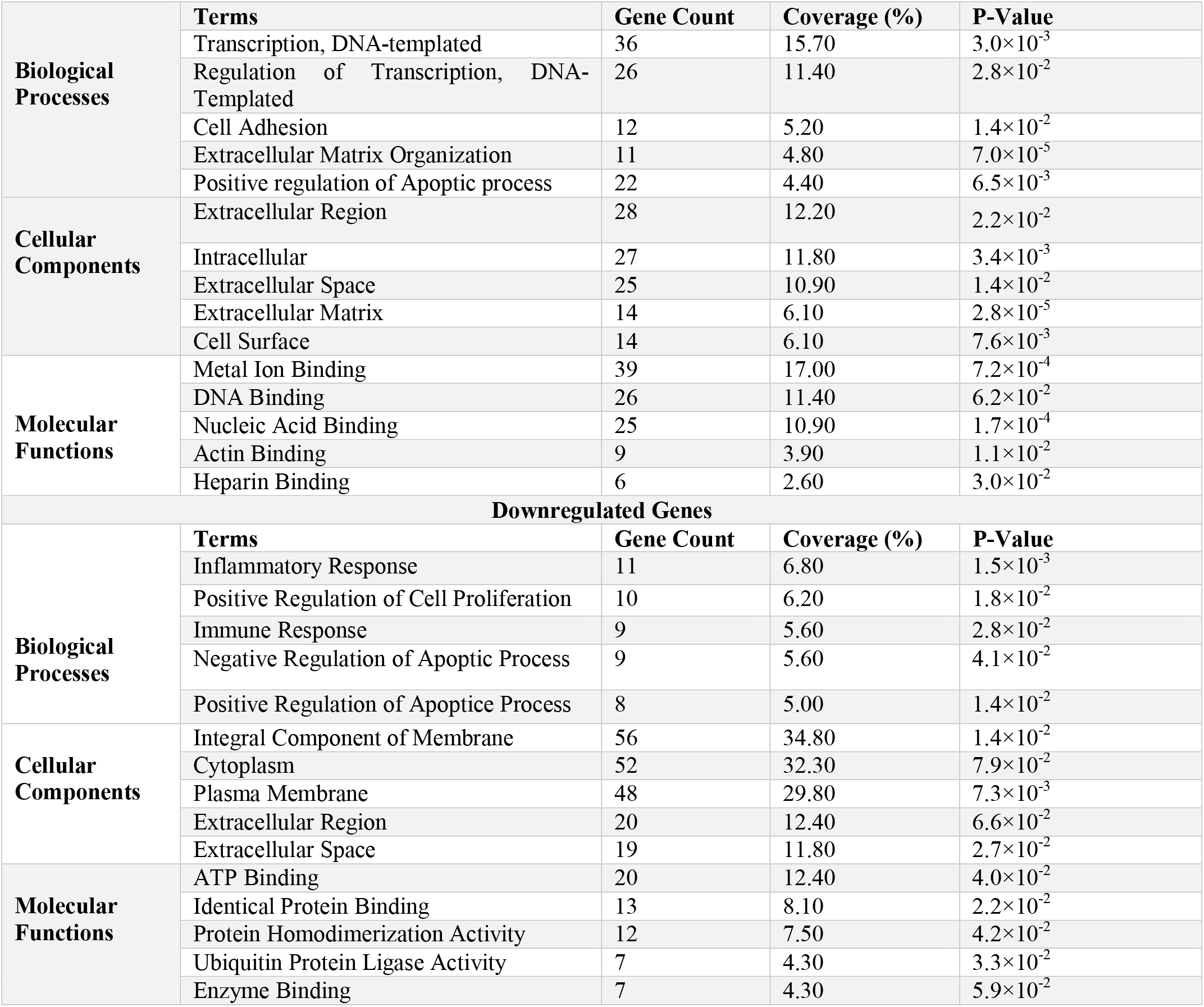
Top five gene ontology (GO) terms of differentially expressed genes (DEGs) in Idiopathic Pulmonary Fibrosis.

**Figure 3:**
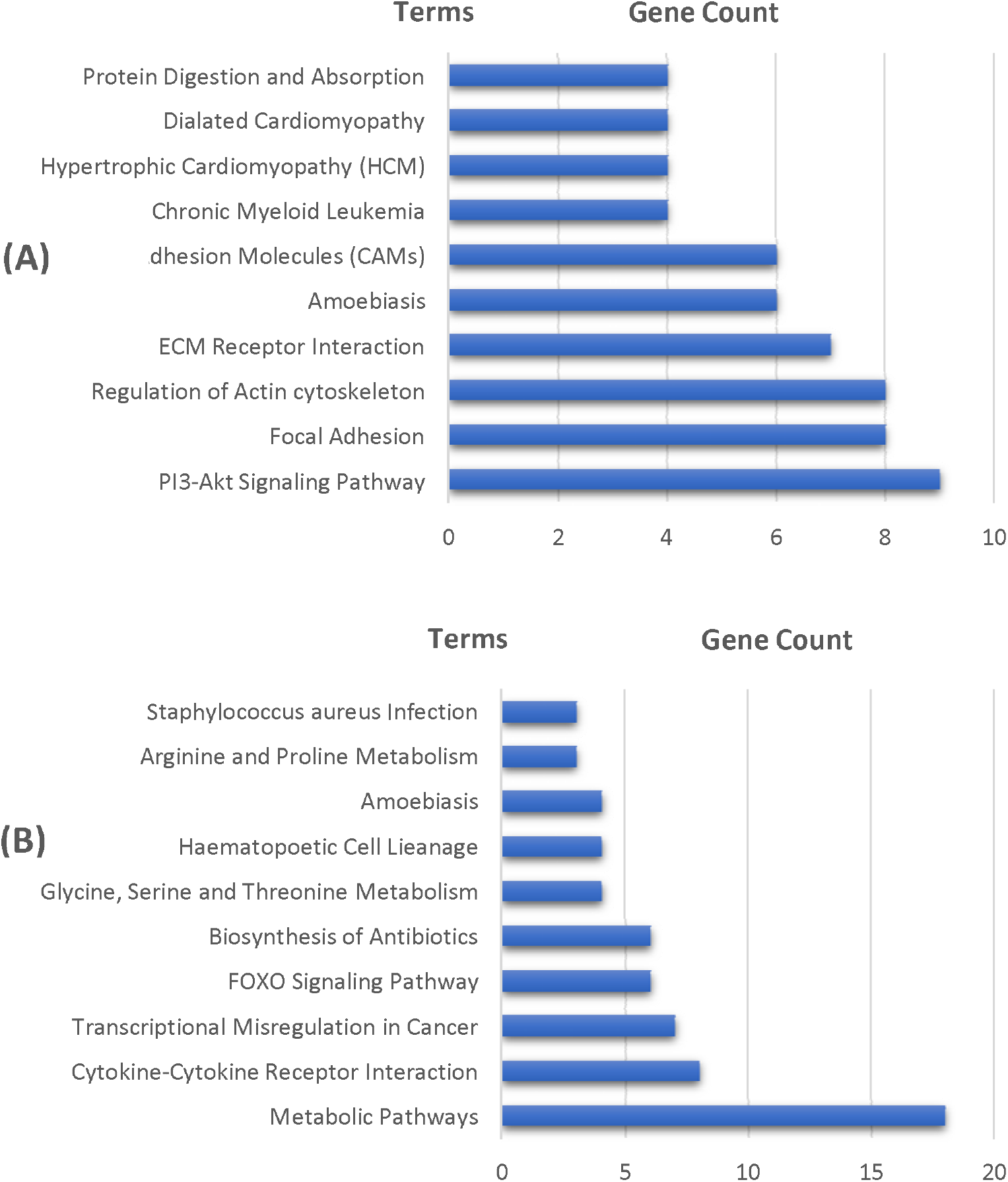
KEGG pathway of differentially expressed genes: (A) Upregulated genes; (B) Downregulated genes.

### 3.2. Proteome Signatures

DEGs were used to construct generic PPI network which generated almost scale free topological densely connected network (**Figure 4**). Then the generic PPI network was utilized in the construction of hub protein network with top 10 most connected nodes (**Figure 5**). UBC, PTEN, SOS1, PTK2, FGFR1, YAP1, FOXO1, RACK1, BMP4 and CD44 were identified as most interacted hub proteins. After that, top 2 modules in the existing network was also analyzed (**Figure 6**). Then the functional enrichment of best 2 modules were also analyzed. Module 1 was mostly involved in protein predominantly in protein acetylation function (P value: 6.86×10^−6^) and module 2 was reported to be involved in G-protein coupled receptor (GPCR) signaling (P value 2.8×10^−4^).

**Figure 4:**
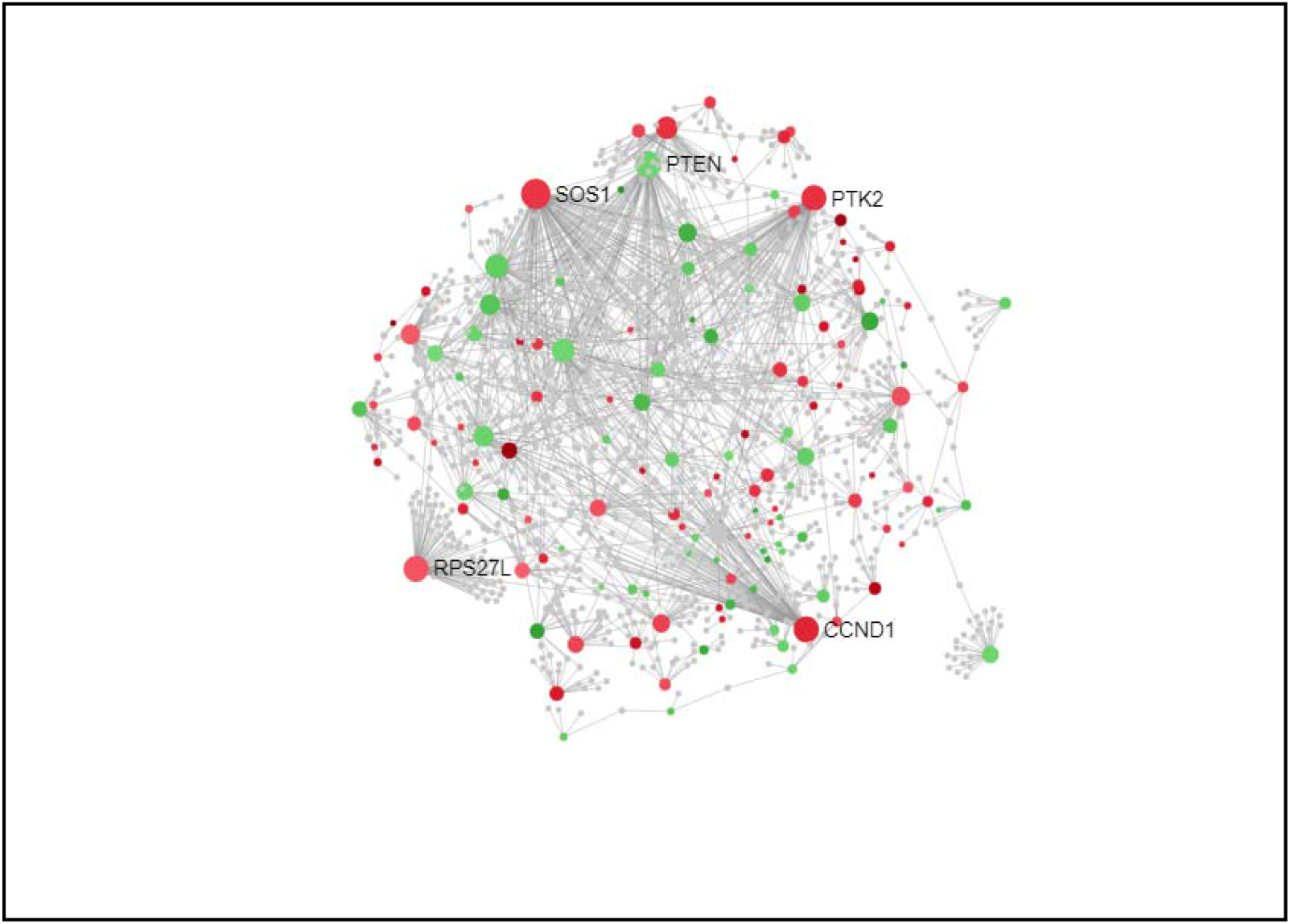
Protein-protein interaction (PPI) network of differentially expressed genes (DEGs). Nodes represent DEGs (Green: Upregulated genes; Red: Downregulated genes). Edges represent interaction.

**Figure 5:**
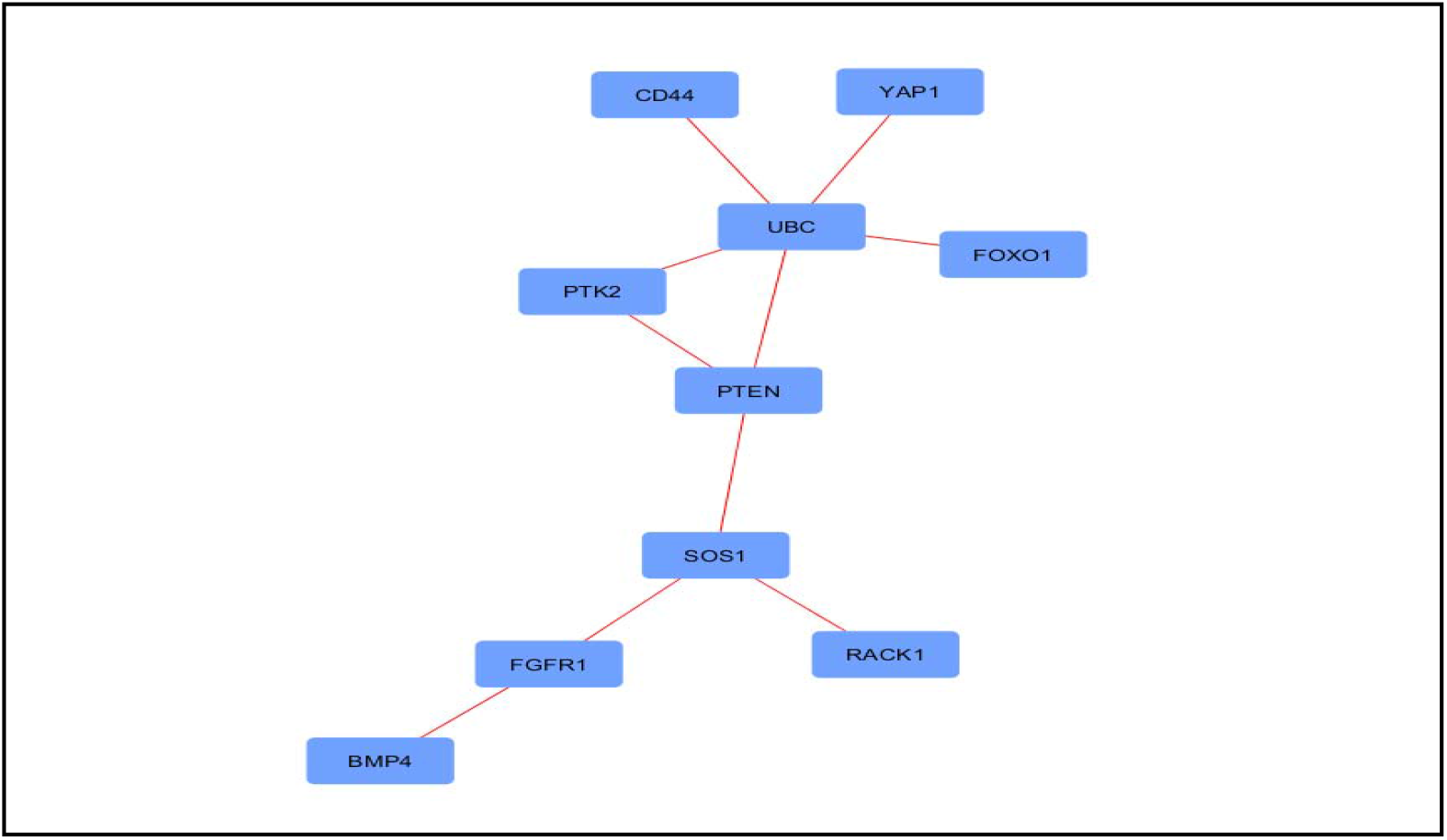
Hub proteins from generated protein-protein interaction (PPI) network. Nodes represent proteins and edges represent interactions.

**Figure 6:**
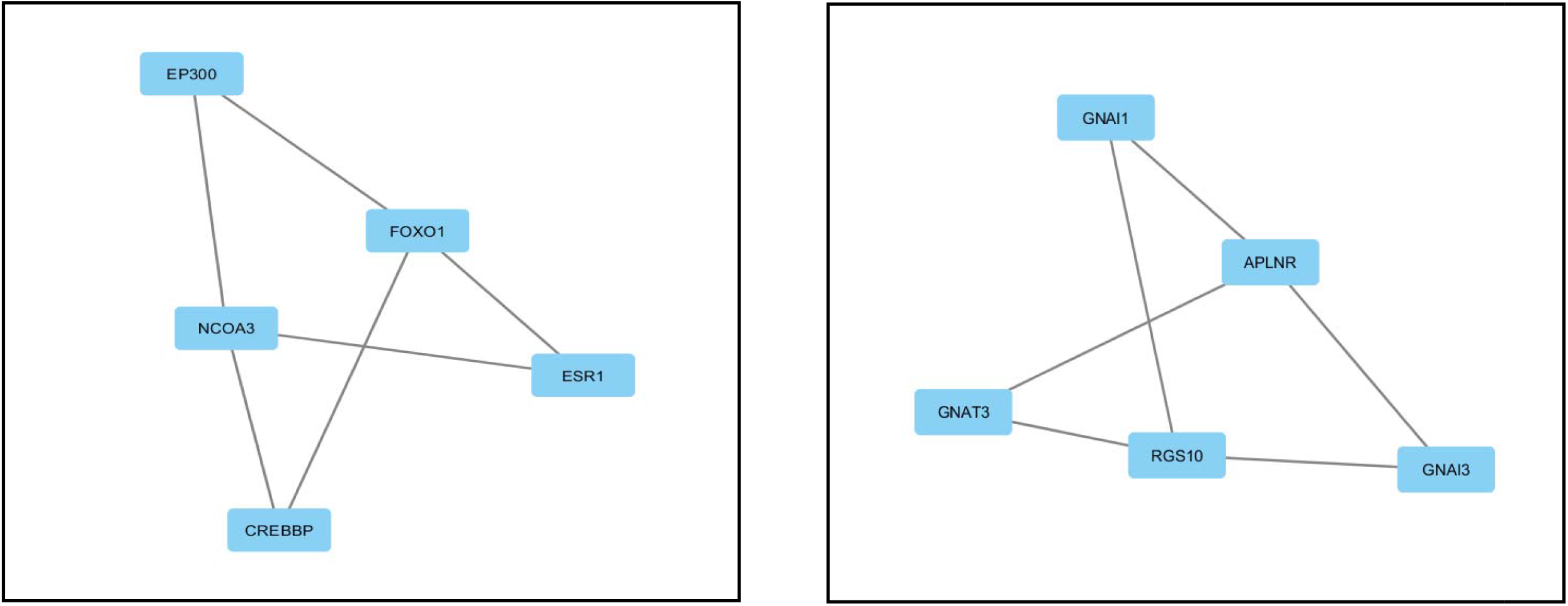
Best two modules obtained from protein-protein interaction (PPI) network in IPF Nodes represent proteins and edges represent interactions

### 3.3. Regulatory Signatures

To identify the regulatory signatures, transcriptional regulatory elements (**Figure 8**) and post-transcriptional regulatory elements (**Figure 9**) were identified from TF/miRNA-DEGs networks. Top 10 transcription factors (E2F1, STAT3, PPARG, MEF2A, FOXC1, GATA3, YY1, GATA2, NFKB1, and FOXL1) and miRNAs (hsa-mir-155-5p, hsa-mir-16-5p, hsa-mir-17-5p, hsa-mir-19a-3p, hsa-mir-192-5p, hsa-mir-92a-3p, hsa-mir-26b-5p, hsa-mir-335-5p, hsa-mir-124-3p, and hsa-let-7b-5p) were selected from the networks as the best signatures.

**Figure 7:**
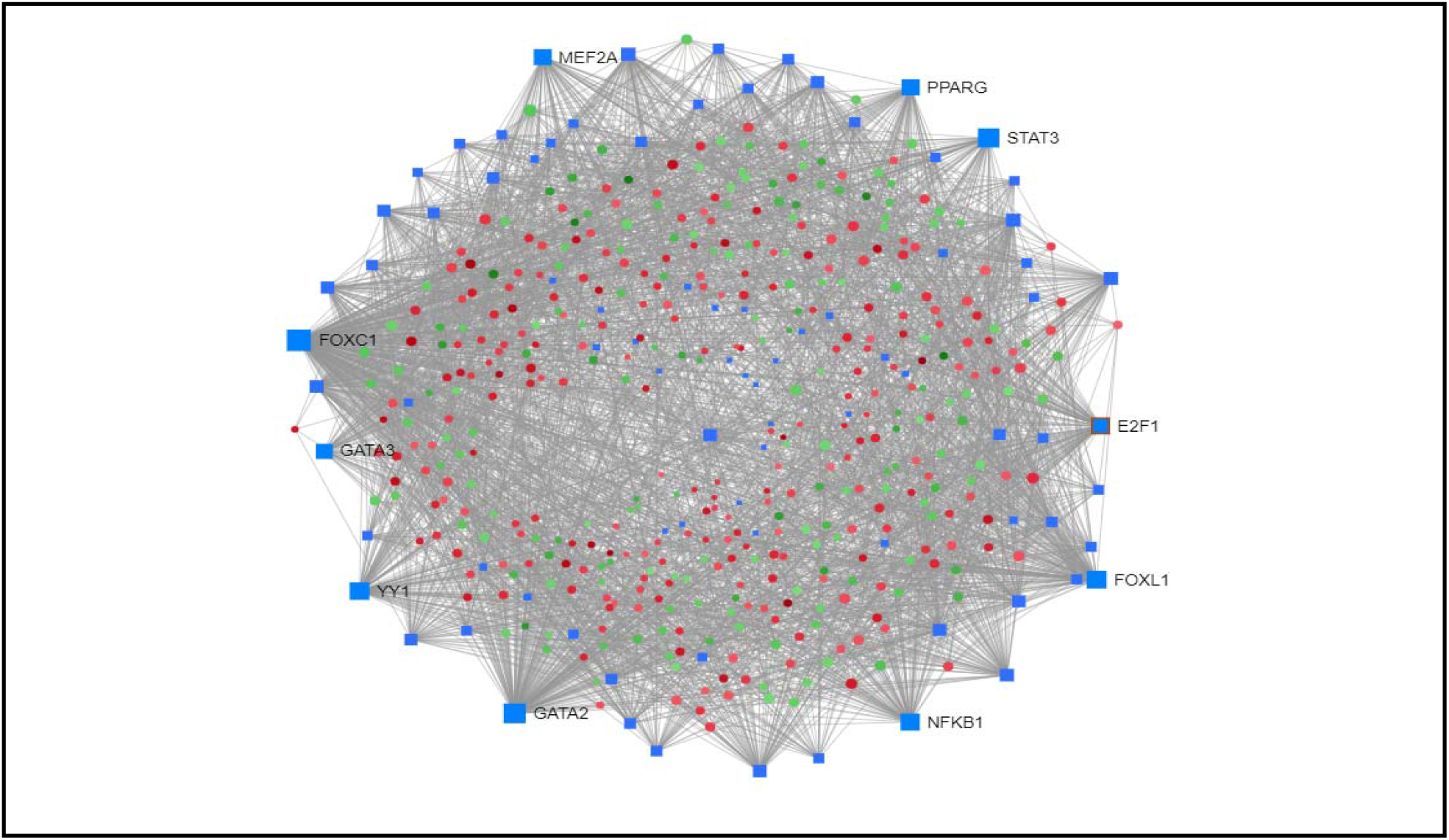
Interaction between transcription factor (TF) and differentially expressed genes network of differentially expressed genes (DEGs). Nodes represent DEGs (Green: Upregulated genes; Red: Downregulated genes; Blue: Transcription factors). Edges represent interaction.

**Figure 8:**
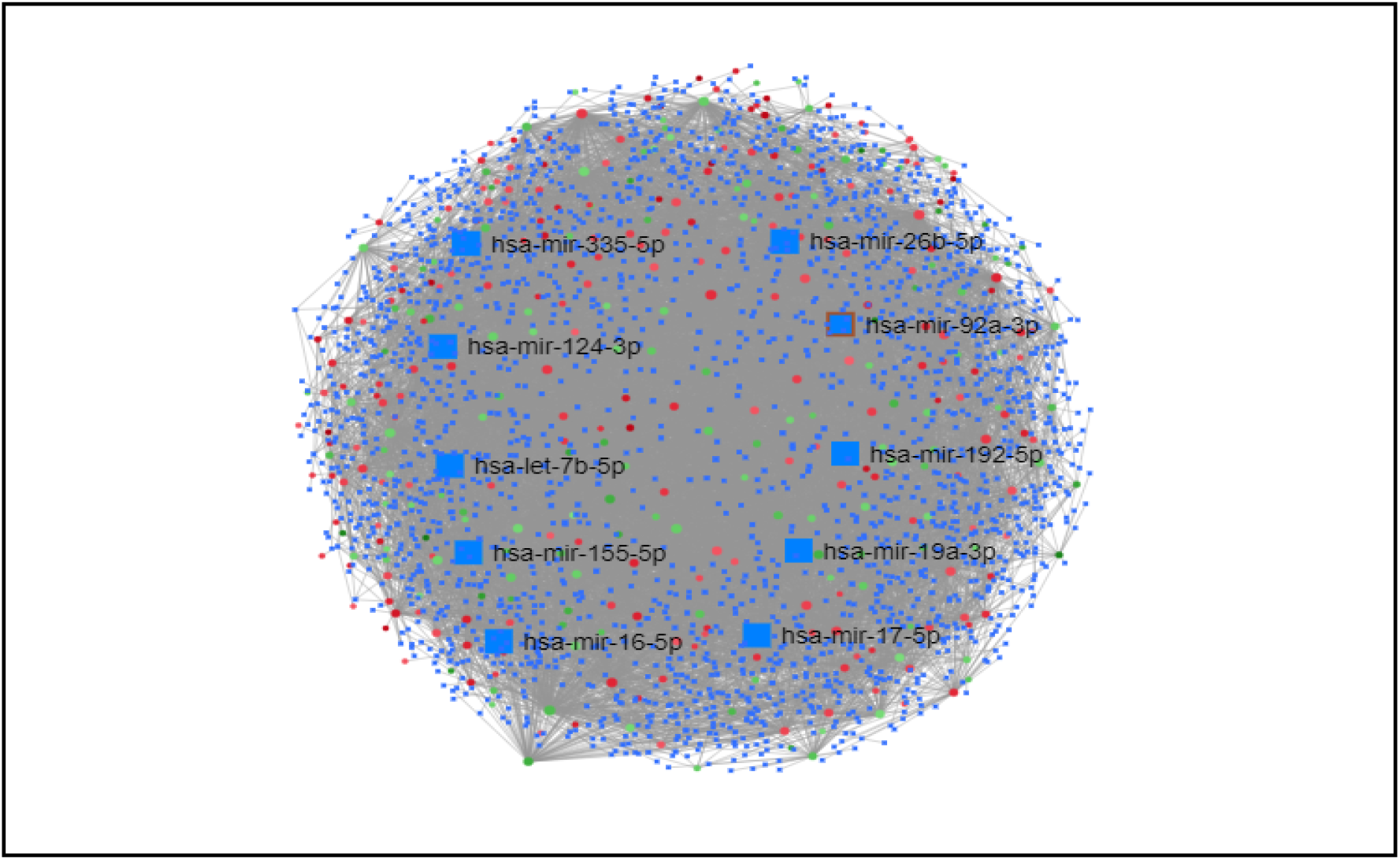
Interaction between miRNA and differentially expressed genes network of differentially expressed genes (DEGs). Nodes represent DEGs (Green: Upregulated genes; Red: Downregulated genes; Blue: miRNA). Edges represent interaction.

## 4. Discussion

In order to identify potential biomarkers or drug target in IPF gene expression pattern was analyzed from array data. 660 differentially expressed genes were identified after statistical analysis. Again, their involvement in different biological processes, cellular compartmentation and molecular functions were also analyzed. The selected DEGs indicated their predominant involvement in transcription, extracellular matrix organization, cell adhesion, apoptotic process and binding activity (**Table 1**). IPF is assumed to be occurred by the manifestation of multiple pathways. A range of cascades including apoptotic pathway, inflammatory cytokines, extracellular matrix regulatory factors, vascular endothelial remodeling have been implicated in animal models of fibrosis (Maher et al., 2007).

Moreover, in order to identify hub proteins and regulatory biomolecules their PPI network and TF/miRNA-DEGs network were also analyzed. Protein-protein interaction is central to understand the functional relationships between different proteins and a protein-protein interaction map provides the functional organization of the proteome of specific interest (Xenariosand Eisenberg, 2001; Stelzl et al., 2005). We utilized BottleNeck method to construct the hub protein interactome. BottleNeck is more accurate in predicting the protein interactome than degree matrix (Yu et al., 2007). Among the top 10 hub proteins, UBC (Ubiquitin-C) mediates proteasome dependent proteolysis, transcriptional regulation, apoptosis and many other functions (Radici et al., 2013). Elevated level of UBC has been reported in patients with Chronic Obstructive Pulmonary Disease (COPD) and ubiquitination and proteolysis have been shown to play a crucial role in COPD (Létuvé et al., 2010; Debigaré et al., 2010). PTEN (Phosphatase and Tensin Homolog) is a cell membrane phosphatase. Decreased PTEN activity has been demonstrated in the cell membrane of IPF fibroblast (Xia et al., 2010). Forkhead Box (FOXO) 1 along with FOXO3 have been shown to have favorable inhibitory effect on fibrosis activation and reduction in extracellular matrix production. These two proteins have already been suggested as potential drug target in fibrosis (Xin et al., 2018; Al□Tamari et al., 2018). The transmembrane glycoprotein CD44 comprises several isoforms and plays crucial role in cell-cell adhesion. A different form of CD44 expression pattern was observed in plexiform lesions of idiopathic pulmonary arterial hypertension (IPAH) (Ohta□Ogo et al., 2012). Yet another study evident different expression patterns of CD44 including more isoforms in fibrotic lung samples (Kasper et al., 1995). Recent study with Yes-Associated Protein 1 (YAP1), a key regulator of Hippo pathway, revealed its overexpression leading to cell proliferation, migration and collagen production in IPF whereas knockdown of YAP1 resulted in reduced fibroblast aggregation with amelioration of the fibrosis condition bot *in vivo* and *in vitro* (Chen et al., 2019). FGFR1 (Fibroblast Growth Factor 1) plays key role in the development of squamous cell lung cancer (Sekine et al., 2014). Upregulation of RACK1 (Receptor of Activated C Kinase 1) has been shown to induce hepatic fibrosis in mice (Jia et al., 2013).

Moreover, two modules (**Figure 6)** were selected from the generic PPI network which was predicted to majorly be involved in protein acetylation and GPCR signaling. Abnormal pattern of histone acetylation has been observed in IPF lung tissue which was evident by low level of COX-2 production in laboratory experiment (Coward et al., 2009). Moreover, abnormal histone deacetylation has also been reported in lung tissues of IPAH patient (Nozik-Grayck et al 2016). Transcriptional signatures i.e., TFs and Post-transcriptional signatures i.e., miRNAs provide potential sources in biomarker identification and drug targeting (Islam et al., 2018). DEGs were mapped to construct TF/miRNA-DEGs interaction network in order to find regulatory biomolecule signatures. Among the top 10 selected TFs (**Figure 7**), GATA3 Binding Protein (GATA3) overexpression in mouse model has been shown to enhance the development of IPF (Kimura et al., 2006). Forkhead Box C1 (FOXC1) encoding gene has been found to be differentially methylated in laboratory experiment with IPF subject (Yang et al., 2014). MEF2A (Myocyte Enhancer Factor 2A) has been reported to influence the growth and proliferation of lung fibroblast (Han et al., 2015). PPARG (Peroxisome Proliferator-Activated Receptor Gamma) is assumed to control the gene expression in lung fibrosis and it has already been suggested as a potential drug target in IPF (Dumoulin et al., 2010). GATA2 (GATA2 Binding Protein) deficiency is a frequent feature in pulmonary diseases. Association of GATA2 deficiency has been evident in both fibrosis and Pulmonary Alveolar Proteinosis (PAP) (Svobodova et al., 2015; Ballerie et al., 2016). STAT3 (Signal Transduce and Activator of Transcription 3) plays multiple crucial roles in maintaining lung homeostasis and it is assumed to be one major target in lung fibrosis (Prêle et al., 2012). YY1 (Yin Yang 1) is a transcriptional repressor which has been suggested as novel regulator of pulmonary fibrosis. Overexpression of YY1 has been reported in both human IPF and murine model and it has been claimed that YY1 promotes fibrogenesis at least in part by increasing collagen or α-SMA secretion (Lin et al., 2011).

Among the top 10 selected miRNAs, hsa-mir-155-5p, hsa-mir-16-5p and hsa-mir-26b-5p have been predicted to be involved in multiple pathways giving rise to interstitial lung diseases (ILD) and suggested as non-invasive biomarker (Mishra et al., 2018), hsa-mir-17-5p and hsa-mir-19a-3p have been shown to be involved in the prognosis of small cell lung cancer (Hayashita et al., 2005; Mancuso et al., 2016). Again, downregulation of hsa-mir-335-5p expression has been reported in COPD patients with PiZZ (Glu342Lys) inherited alpha1-antitrypsin deficiency (AATD) (Esquinas et al., 2017). In yet other laboratory experiments, hsa-mir-124-3p has been shown to participate in pulmonary vascular remodeling in association with a long non-coding RNA (lncRNA) called MALAT1 (Metastasis Associated Lung Adenocarcinoma Transcript 1), and hsa-let-7b-5p has been shown to be downregulated in patients with cystic fibrosis (CF) (Wang et al., 2019; Ideozu et al., 2019).

Finally, the identified hub proteins and regulatory biomolecules may provide potentials sources of non-invasive biomarkers which could be used in targeting drugs against IPF and early diagnosis of this disease. *In vit*ro and *in vivo* experimental evidences also suggested their potential roles and connections with IPF and other related lung diseases. This study suggests (UBC, PTEN, SOS1, PTK2, FGFR1, YAP1, FOXO1, RACK1, BMP4 and CD44) as the best proteome signatures, (E2F1, STAT3, PPARG, MEF2A, FOXC1, GATA3, YY1, GATA2, NFKB1, and FOXL1) as the best transcription regulatory signatures, and (hsa-mir-155-5p, hsa-mir-16-5p, hsa-mir-17-5p, hsa-mir-19a-3p, hsa-mir-192-5p, hsa-mir-92a-3p, hsa-mir-26b-5p, hsa-mir-335-5p, hsa-mir-124-3p, and hsa-let-7b-5p) as the best post-transcription regulatory signatures in IPF within the selected strategies employed in the exploration of biomarkers.

## 5. Conclusion

The underlying mechanism of IPF is poorly understood and this is the major hindrance to develop effective drug and early diagnosis method. In this study network-based approach has been utilized to analyze microarray data in order to identify potential transcriptome, proteome and regulatory signatures. Different hub genes involved in apoptic process, transcription, inflammatory responses, extracellular matrix organization have been identified to play key role. Thereafter multiple hub proteins, transcription factors and micro RNAs playing crucial roles in IPF and other related lung diseases were also identified and these findings were supported by other available laboratory experimental data. We suggest further exploration and possible laboratory experiments with the identified signatures to confirm their superiority in the diagnosis and drug targeting against IPF. Hopefully, this study will raise research interest among researchers and contribute in the discovery of non-invasive and effective biomarkers and potential drug targets.

## Data Availability

Authors made all data generated during experiment and analysis available in the manuscript.

## Acknowledgement

Authors are thankful to the members of Swift Integrity Computational Lab, Dhaka, Bangladesh.

## Data Availability Statement

All the data generated and analyzed are summarized in this manuscript.

